# Basic and effective reproduction numbers of COVID-19 cases in South Korea excluding Sincheonji cases

**DOI:** 10.1101/2020.03.19.20039347

**Authors:** Jeongeun Hwang, Hyunjung Park, Jiwon Jung, Sung-Han Kim, Namkug Kim

## Abstract

In February and March 2020, COVID-19 epidemic in South Korea met a large black swan effect by a Sincheonji cult mass infection in Daegu-Gyeongbuk area. The black swan made it difficult to evaluate that the current policies for infection prevention including social distancing, closing schools, hand washing, and wearing masks good enough or not. Therefore, in this study, we evaluated basic reproduction number (R_0_) and time-dependent reproduction number (R_t_) of confirmed cases based on various kinds of populations, including total, Daegu-Geyoengbuk, except-Daegu-Gyeongbuk, Sincheonji, and except-Sincheonji. In total, it seems the infection is going to be under control, but this is never true because in the except-Sincheonji and except-Daegu-Geyongbuk cases, R_0_ is still above 1.0, and R_t_ is drifting around 1.0. This study could be used to determine government policies in the near future.

## Text

South Korea currently has 9,241 cumulated confirmed COVID-19 cases as of March 26^th^, 2020. There was a large “black swan” event called the Sincheonji cult mass infection incidence that lead to an exponential increase of confirmed cases. Since then, a great effort has been made to defend the public from virus, and to some extent, these efforts have suppressed the spread.

The basic and time-dependent effective reproduction number R_0_ and R_t_ are two of the most representative characteristics for the dynamics of the infectious virus. To observe the basic features of the COVID-19 outbreak in South Korea without the black swan effect, we collected the daily counts of confirmed cases. The Korean Centers for Disease Control and Prevention (KCDC) has investigated, summarized, and released daily updates on the cumulated counts of confirmed cases, which are available online^1^. The cumulated counts have been provided in total, according to the administrative provinces (Gwangyeok-Si and Do in the South Korean administrative system), and several mass infection cases have occurred. We collected counts of daily confirmed cases from the Sincheonji cult, which was named after a church with a COVID-19 outbreak, and subtracted them from the total counts to comprise the “except Sincheonji” cases. Additionally, we collected the case counts from Daegu-si and Gyeongsangbuk-do province, which surrounds Daegu-si, because the initial outbreak in the Sincheonji cult took place in those areas. We summed up the counts from the two provinces and subtracted them from the total counts to total the “except Daegu-Gyeonbuk” cases. The Sincheonji cult cases and Daegu-Gyeongbuk cases are not mutually inclusive nor exclusive. Because it took days to identify a confirmed case belonging to the Sincheonji cult, the time frames of the Sincheonji cases could have been confounded.

The cumulated counts from January 21^st^, 2020 (the first confirmed case of COVID-19 in South Korea) to March 26^th^, 2020 are shown in Figure 1. The first Sincheonji cult case appeared on February 18^th^, 2020, and it was the first case in the Daegu-Gyeongbuk area. Qualitatively, the cumulated counts curves for Daegu-Gyeongbuk and Sincheonji looks like they are reaching for a plateau, but the curves for the except-Sincheonji and except-Daegu-Gyeongbuk cases do not.

**Figure 1.**
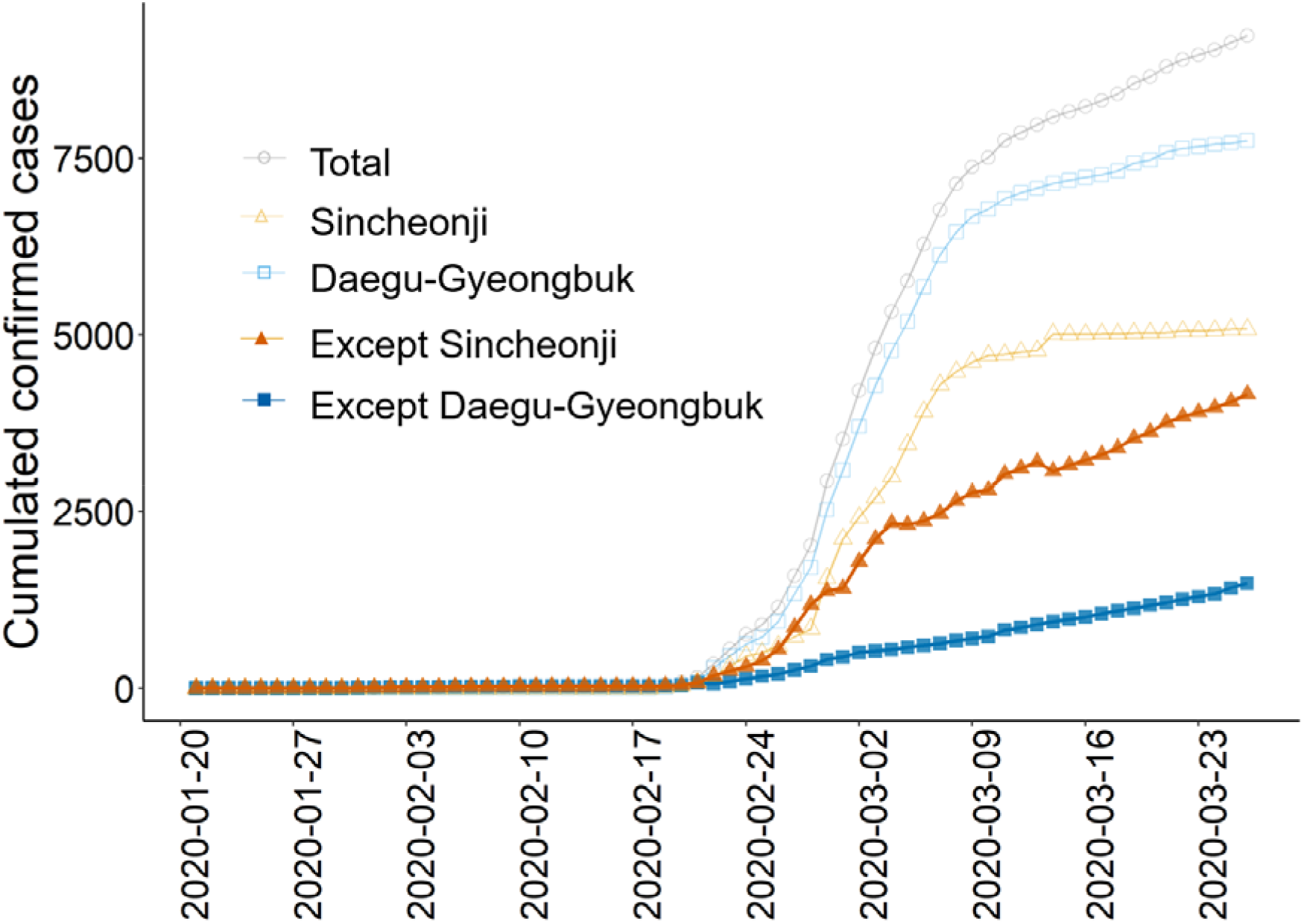
Cumulated counts of confirmed cases of the subgroups, including the total, Daegu-Geyoengbuk, except-Daegu-Gyeongbuk, Sincheonji, and except-Sincheonji cases.

R statistics software version 3.6.3 (R Foundation for Statistical Computing, Vienna, Austria) and the R0 package^2^ was used. For the estimation of the subgroups’ R_0_ values, we fitted an exponential growth model^3^. Sensitivity analysis of the exponential growth period was performed. The generation time distribution should be obtained from the time lag between all infector–infectee pairs^4^, which is called a serial interval, but it is currently unavailable. Instead, we assumed that the serial interval had a gamma distribution with mean ± standard deviation of 2.0 ± 1.0, 3.0 ± 1.5, 4.0 ± 2.0, 5.0 ± 2.5, and 6.0 ± 3.0 days. The serial interval could be approximated by referring to the estimated incubation time of COVID-19. Recent research by Guan et al.^5^ suggested that the virus has an incubation period of four days with an interquartile range two to seven days. Some studies have estimated a wider range for the incubation period; data for human infection with other coronaviruses (e.g. MERS-CoV, SARS-CoV) suggested that the incubation period may range from two to 14 days^6^. Sensitivity tests for the generation time assumptions were performed (Figure S1).

Figure 2 shows the estimated R0 for each study subgroup and their 95% confidence intervals (by simulation) with an assumption of 4.0 ± 2.0 (mean ± standard deviation). In the sensitivity test, the R_0_ values were sensitive to the generation time assumptions; therefore, the numbers themselves are not reliable in the current study. On the contrary, the relative scales among the R_0_ values were robust to the generation time assumptions.

**Figure 2.**
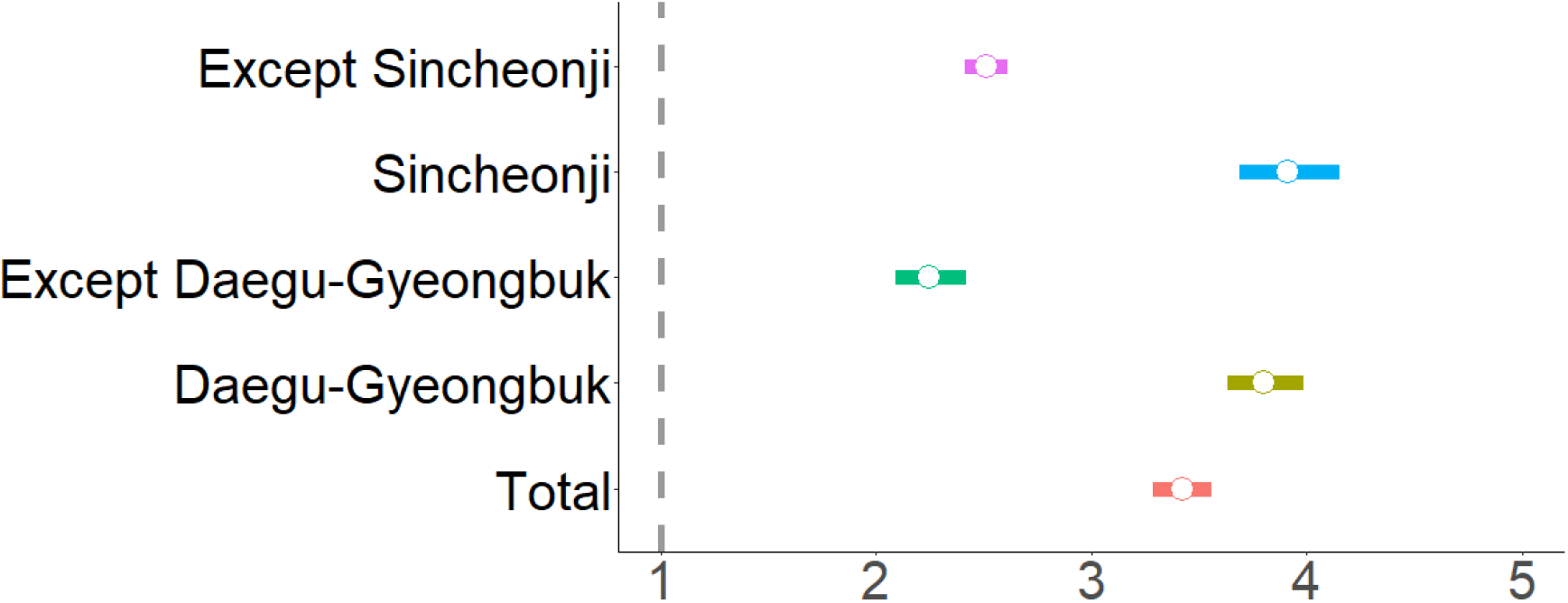
R_0_ with 95% confidence intervals in the subgroups, including the total, Daegu-Gyeongbuk, except Deagu-Gyeongbuk, Sincheonji, and except Sincheonji groups.

The Daegu-Gyeongbuk cases showed significantly higher R_0_ than the except-Daegu-Gyeongbuk cases, and the Sincheonji cases showed significantly higher R_0_ than the except-Sincheonji cases. In all subgroups, R_0_ was larger than 1.0. R_0_ is an initial-stage characteristic before the health authorities take action or social awareness rises. Since the Sincheonji cult mass infection incidence was noticed, health authority actions including COVID-19 tests, isolation, active surveillance, and school closures, were executed immediately. In addition, social awareness of COVID-19 soared, and South Korean residents began to more actively follow hygiene recommendations, including wearing masks, washing hands frequently, and practicing social distancing in everyday life, throughout the nation.

Adding to such efforts to fight the disease, we measured the time dependent reproduction rate R_t_^7^ for each subgroup. We again assumed the generation time to gamma distribution with a 4.0 ± 2.0 mean ± standard deviation.

In Figure 3, R_t_ with 95% confidence intervals in the subgroups, including the total, Sincheonji, except-Sincheonji, Daegu-Gyeongbuk, and except-Daegu-Gyeongbuk subgroups, are shown. In all subgroups, R_t_ gradually decreased until the end of February. R_t_ for the Sincheonji and Daegu-Gyeongbuk cases are staying under R_t_ = 1.0 horizon, but in the except-Sincheonji and except-Daegu-Gyeongbuk cases, R_t_ is drifting around the R_t_ = 1.0 horizon currently (as of March 26^th^, 2020).

**Figure 3.**
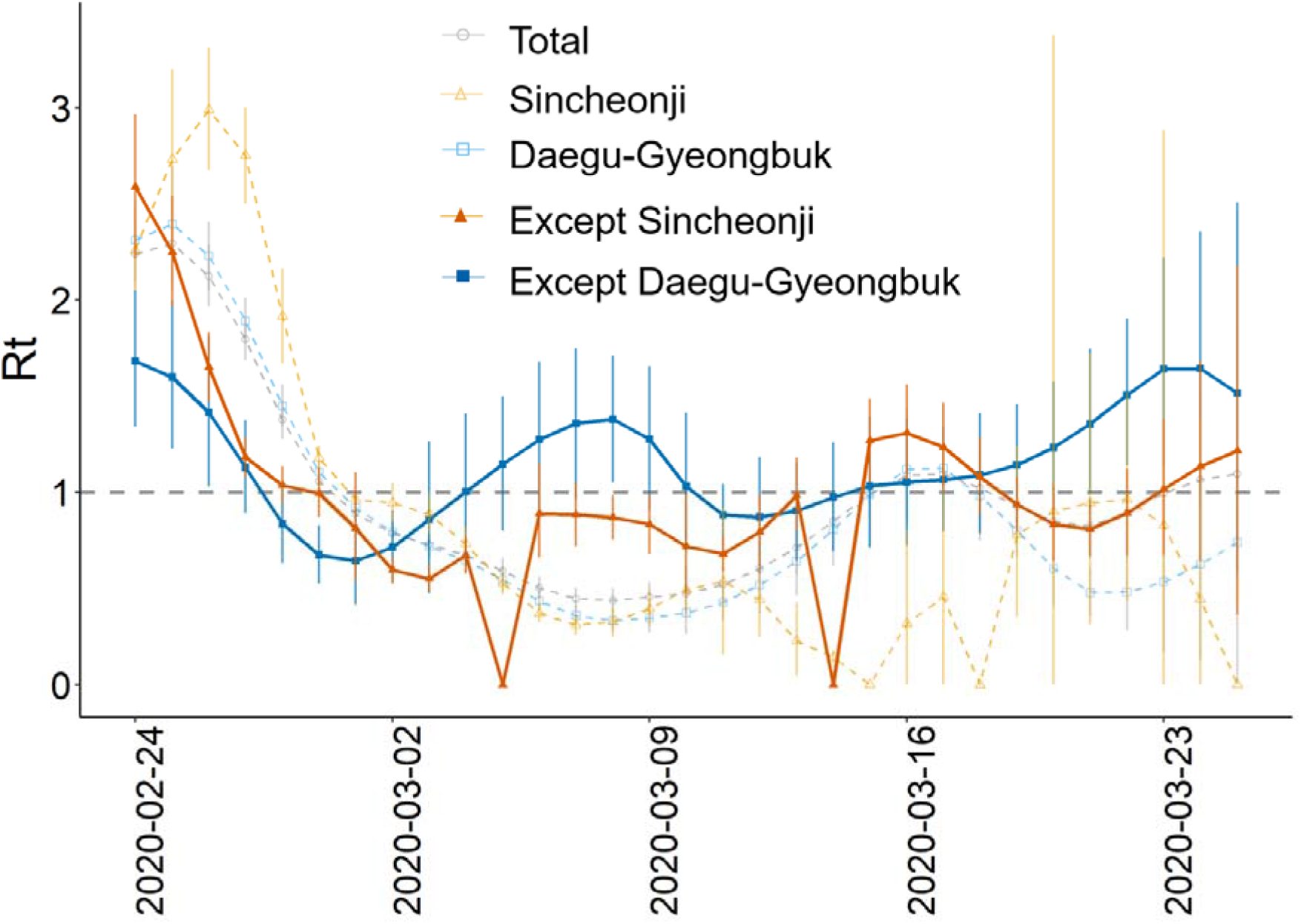
Changes of R_t_ with 95% confidence intervals of subgroups, including the total, Sincheonji, except-Sincheonji, Daegu-Gyeongbuk, and except-Daegu-Gyeongbuk groups. A week after the first Sinceonji cult mass infection case (February 24^th^, 2020), R_t_ for all subgroups decreased for a couple of days. In the following weeks (around March 2^nd^, 2020 and after) Sincheonji and Daegu-Gyeongbuk cases fell under the R_t_ = 1.0 horizon, but R_t_ for the except-Sincheonji and except-Daegu-Gyeongbuk cases were drifting around the 1.0 horizon.

Although the analysis in Figure 3 is preliminary, and thus, we should be very cautious about its interpretations, we see a great uncertainty that South Korea is still in danger of COVID-19. Another instance of the mass infection case could happen if we lose the current momentum in social distancing and preventive action, even if no black swan comes. Assessing only the total counts may blind our sense of danger. Therefore, analyzing the except-Sincheonji and except-Daegu-Gyeongbuk cases separately from the total cases would be recommended. We are releasing the R_t_ estimates’ daily updates for various regional subgroups of South Korea online; http://covid19.mi2rl.co/

Our current analysis has substantial limitations. Because we collected the daily press releases from the KCDC, there could be re-classified cases not reflected in our analysis. There could also be misclassification, because a Sincheonji cult may deny his or her beliefs in the KCDC investigations. Imported cases were not accounted for in the current analysis, though the cumulated imported cases until March 17^th^, 2020 was 54, according to KCDC press release. Another source of confounding is that we are not aware of the true number of cases. In a recent study in China, Li et al.^8^ estimated that only 14% of infected cases have been confirmed by the authorities. Nevertheless, qualitative interpretations may stay robust to under-reporting if they are constant. Delays in confirmation and in classification are another source of error. The uncertainty in the generation time is also an issue, but it could be overcome by performing sensitivity analysis. If more detailed data on infector–infectee interactions becomes available, more reliable analysis could be achieved.

To conclude, in total, it seems that the virus is going to be under control, but this is never true because if we analyze the except-Sincheonji and except-Daegu-Geyongbuk cases, the basic reproduction rate is still above 1.0, and the effective reproduction rate Rt is still higher than 1.0. The current efforts to suppress the COVID-19 outbreak should at least be maintained.

## Data Availability

We used public data only

## Author Contributions

Conceptualization: Hwang J, Kim S, Jung J, Kim N. Methodology: Hwang J, Kim N. Formal analysis: Hwang J, Kim N. Data curation: Par H, Software: Hwang J, Kim N. Investigation: Jung J, Kim S, Kim N. Writing – original draft preparation: Hwang J, Park H. Writing – review and editing: Jung J, Kim S, Kim N. Approval of final manuscript: all authors.

## REFERENCES

1. KCDC. Korean Centers for Disease Control and Prevention http://cdc.go.kr 2020.

2. Obadia T, Haneef R, Boelle PY. The R0 package: a toolbox to estimate reproduction numbers for epidemic outbreaks. Bmc Medical Informatics and Decision Making 2012;12.

3. Wallinga J, Lipsitch M. How generation intervals shape the relationship between growth rates and reproductive numbers. Proceedings of the Royal Society B-Biological Sciences 2007;274(1609):599–604.

4. Svensson A. A note on generation times in epidemic models. Mathematical Biosciences 2007;208(1):300–11.

5. Guan WJ, Ni ZY, Hu Y, Liang WH, Ou CQ, He JX, et al. Clinical Characteristics of Coronavirus Disease 2019 in China. N Engl J Med 2020.

6. CDC. Interim Clinical Guidance for Management of Patients with Confirmed Coronavirus Disease (COVID-19). https://www.cdc.gov/coronavirus/2019-ncov/hcp/clinical-guidance-management-patients.html. Updated March 7, 2020 2020.

7. Wallinga J, Teunis P. Different epidemic curves for severe acute respiratory syndrome reveal similar impacts of control measures. American Journal of Epidemiology 2004;160(6):509–16.

8. Li R, Pei S, Chen B, Song Y, Zhang T, Yang W, et al. Substantial undocumented infection facilitates the rapid dissemination of novel coronavirus (SARS-CoV2). Science 2020.

